# Randomized Controlled Trials Evaluating AI in Clinical Practice: A Scoping Evaluation

**DOI:** 10.1101/2023.09.12.23295381

**Authors:** Ryan Han, Julián N. Acosta, Zahra Shakeri, John P.A. Ioannidis, Eric J. Topol, Pranav Rajpurkar

## Abstract

**Background:** Artificial intelligence (AI) has emerged as a promising tool in healthcare, with numerous studies indicating its potential to perform as well or better than clinicians. However, a considerable portion of these AI models have only been tested retrospectively, raising concerns about their true effectiveness and potential risks in real-world clinical settings.

**Methods:** We conducted a systematic search for randomized controlled trials (RCTs) involving AI algorithms used in various clinical practice fields and locations, published between January 1, 2018, and August 18, 2023. Our study included 84 trials and focused specifically on evaluating intervention characteristics, study endpoints, and trial outcomes, including the potential of AI to improve care management, patient behavior and symptoms, and clinical decision-making.

**Results:** Our analysis revealed that 82·1% (69/84) of trials reported positive results for their primary endpoint, highlighting AI’s potential to enhance various aspects of healthcare. Trials predominantly evaluated deep learning systems for medical imaging and were conducted in single-center settings. The US and China had the most trials, with gastroenterology being the most common field of study. However, we also identified areas requiring further research, such as multi-center trials and diverse outcome measures, to better understand AI’s true impact and limitations in healthcare.

**Conclusion:** The existing landscape of RCTs on AI in clinical practice demonstrates an expanding interest in applying AI across a range of fields and locations. While most trials report positive outcomes, more comprehensive research, including multi-center trials and diverse outcome measures, is essential to fully understand AI’s impact and limitations in healthcare.

## INTRODUCTION

The use of artificial intelligence (AI) in healthcare has seen significant growth in recent years, with several publications reporting that medical AI models can perform as well or better than clinicians.^1–3^ However, many of these models have only been tested retrospectively, using surrogate endpoints, and outside of real-life clinical settings. Out of nearly 300 AI-enabled medical devices approved or cleared by the FDA, only a few have undergone evaluation using prospective, randomized controlled trials.^4^

The lack of real-world evaluation of AI systems leaves substantial uncertainty, even for the possibility of significant risk to patients and clinicians. One example of this is a widely used sepsis model that was found to have “substantially worse” performance than was reported by its developer, leading to “a large burden of alert fatigue” due to incorrect or irrelevant alerts.^5^ It may not be uncommon for AI to perform worse when deployed prospectively, and the difficulty of adopting AI in a clinical setting can further decrease any potential benefits for outcomes that matter.^6,7^

To provide a clearer understanding of the AI landscape in healthcare, this scoping evaluation examines the state of randomized controlled trials (RCTs) for AI algorithms being used in clinical practice. We focus on evaluating intervention characteristics, study endpoints, and trial outcomes across various fields and locations. Our analysis delves into the potential of AI to improve care management, patient behavior and symptoms, and clinical decision-making, while also identifying areas that require further research. By doing so, we aim to help stakeholders better comprehend the clinical relevance of AI and guide future research in this rapidly evolving domain.

## METHODS

### Search strategy and selection criteria

We conducted a systematic literature search using PubMed and the International Clinical Trials Registry Platform (ICTRP) to identify studies published between January 1, 2018, and August 18, 2023. Our search strategy used free text-terms related to “artificial intelligence”, “clinician”, and “clinical trial”, and is available in the Appendix. We also manually reviewed the references of relevant publications to identify additional articles.

We included randomized controlled trials (RCTs) that met the following criteria: (1) the intervention had a significant component of AI, defined as a non-linear computational model; (2) the intervention was integrated into clinical practice, impacting the management of a patient’s health by a clinical team; and (3) results were published as a full-text article in a peer-reviewed English-language journal. We excluded studies evaluating linear risk scores, secondary studies, abstracts, and interventions that are not integrated into clinical practice. The protocol was registered with PROSPERO (CRD42022326955).

### Data analysis

To ensure the quality of our search results, we utilized the Covidence Review software to screen titles and abstracts. Two independent investigators (RH and JNA) conducted the initial screening, followed by a full-text review of all identified papers. Any discrepancies were resolved through discussion with a third reviewer (PR). Eligible papers were then subjected to data extraction by two reviewers in Google Sheets.

We extracted study-level information, including study location, clinical task, primary endpoint, comparator, and result, as well as the type and origin of the AI used. Additionally, we classified studies by primary endpoint group (diagnostic accuracy, clinical decision making, patient behavior and symptoms, care management), clinical area or specialty, and data modality used by the AI.

We did not attempt to contact study authors for additional or uncertain information. Due to the expected heterogeneity in tasks and endpoints, we did not conduct formal meta-analyses. Instead, we present simple descriptive statistics to provide an overview of the features of the eligible trials.

## RESULTS

Our electronic search retrieved 4,825 study records and 4,079 trial registrations resulting in 8,892 records overall after deduplication. After title and abstract screening, 124 articles were retained for full-text review. Of these, 53 were excluded, leaving 71 studies after the primary screening. An additional 13 articles were identified through secondary reference screening, resulting in a total of 84 unique RCTs included in our study. The references and characteristics for all the included studies are available in the appendix.

**Figure 1.**
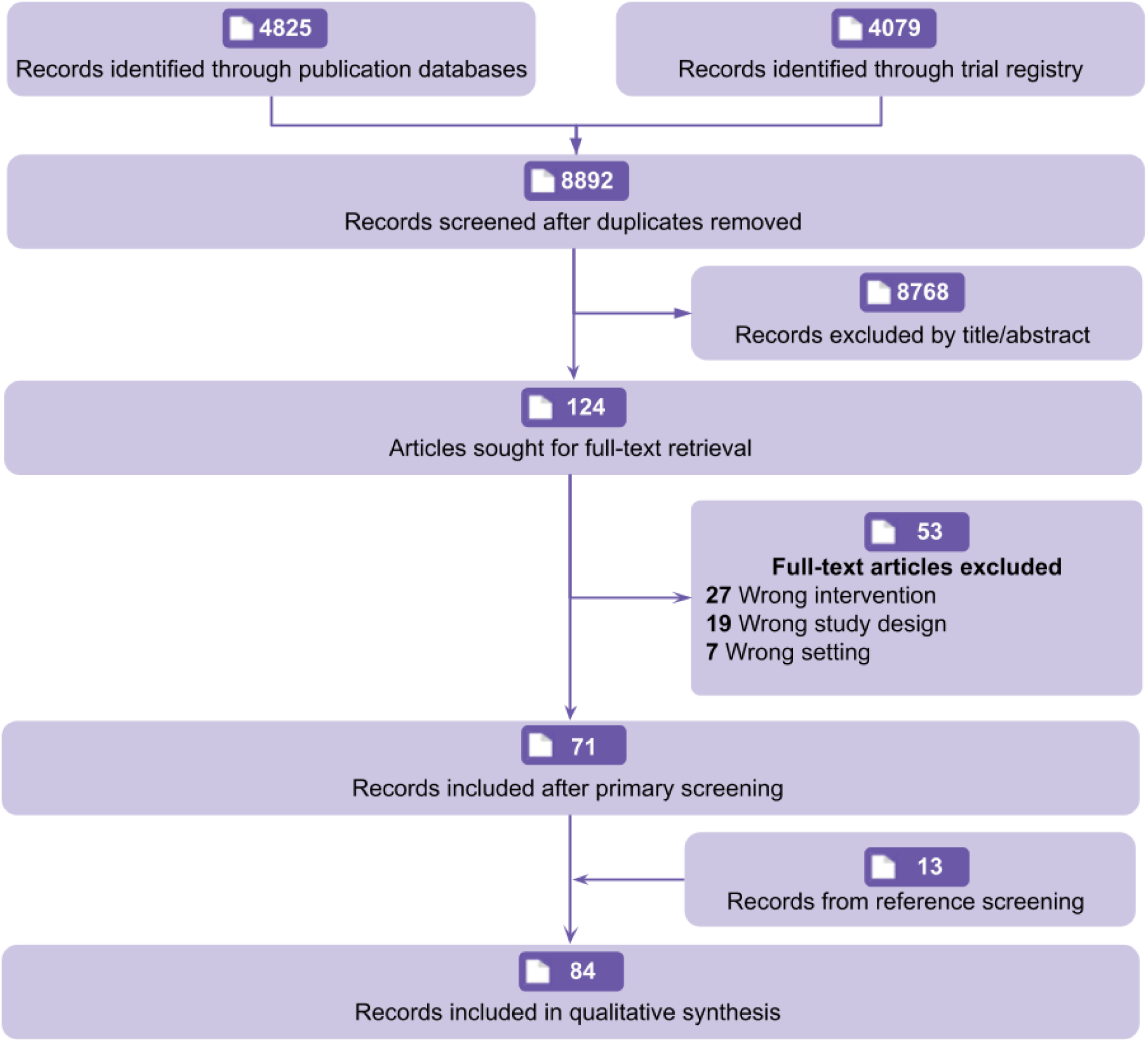
PRISMA (preferred reporting items for systematic reviews and meta-analyses) flowchart of study records.

### In what medical specialties and countries are trials being done?

Many RCTs were related to gastroenterology (35/84, 41·7%), followed by radiology (13/84, 15·5%), surgery (5/84, 6·0%), and cardiology (5/84, 6·0%). Gastroenterology trials were notable for their uniformity, with all trials testing video-based deep learning algorithms in an assistive setup with clinicians, and all but one trial measuring a primary endpoint relating to diagnostic accuracy (detection rate, miss rate, etc.). The majority of gastroenterology trials (24/35, 68·6%) were conducted by only four groups (eight trials from Wuhan University, six trials from Wision AI, six trials from Medtronic, and four trials from Fujifilm).

Most RCTs were conducted in a single country (77/84, 91·7%), with the US (26/84, 31·0%) having the most trials followed by China (24/84, 28·6%). Trials conducted in the US were distributed across various specialties, whereas trials conducted in China predominantly related to gastroenterology (19/24, 79·2%). Trials were predominantly conducted in a single center (52/84, 61·9%) and included a median of 359 patients (Q1 - Q3: 146 - 1054) in their final analysis. Trials conducted in multiple countries primarily involved European nations.

**Figure 2.**
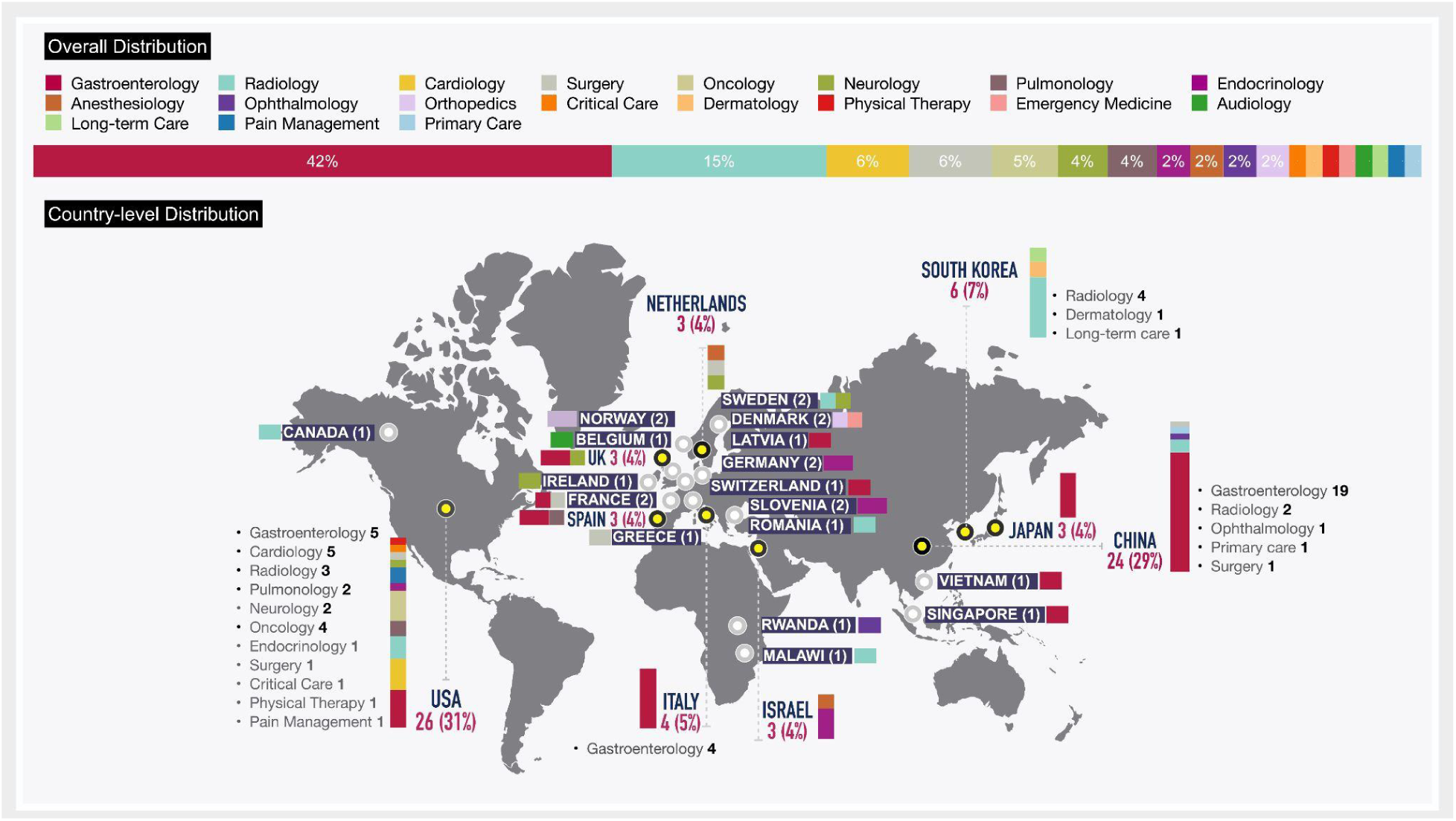
Distribution of randomized controlled trials of AI in clinical practice across countries and specialties.

### What are the outcomes assessed?

Approximately half of the trials (46/84, 54·8%) had primary endpoints relating to diagnostic accuracy, such as detection rate or mean absolute error. Other primary endpoints were grouped according to care management (16/84, 19·0%), patient behavior and symptoms (15/84, 17·9%), and clinical decision-making (7/84, 8·3%). Table 1 summarizes the distribution of results and endpoint types.

**Table 1.** Tabulated summary of primary endpoint results and primary endpoint types for randomized controlled trials of AI in clinical practice.

|  | Primary Result |  |  |  |  |
| --- | --- | --- | --- | --- | --- |
| Comparison | Significant improvement | No significant improvement | Demonstrated noninferiority | Significant deterioration | Grand Total |
| Care management | 13 | 1 | 2 | .. | 16 |
| Clinical decision making | 6 | 1 | .. | .. | 7 |
| Diagnostic accuracy | 35 | 9 | 1 | 1 | 46 |
| Patient behavior and symptoms | 10 | 3 | 2 | .. | 15 |
| Grand Total | 64 | 14 | 5 | 1 | 84 |

A number of RCTs have assessed the impact of AI interventions on care management quality metrics, providing an outcome-oriented view of the use of AI in clinical practice. For example, AI systems for insulin dosing and hypotension monitoring have been shown to improve the average time that patients spend within target ranges for glucose and blood pressure, respectively.^8–11^ Similarly, trials assessing AI systems for radiation therapy and prostate brachytherapy have been evaluated by their ability to reduce rates of acute care and the volume of the prostate tumor.^12,13^

AI systems have also been evaluated in terms of their impact on patient behavior and symptoms. For instance, one trial reported that making AI predictions for diabetic retinopathy risk immediately available to patients increased referral adherence compared to having patients wait for grading by clinicians.^14^ Another trial reported that the adoption of a nociception monitoring system was able to decrease postoperative pain scores in patients relative to unassisted clinicians.^15^ These trials highlight the potential for AI interventions to have a direct impact on patient experience.

Trials have also measured the ability of AI systems to influence clinical decision-making. For example, the availability of AI mortality predictions for cancer patients was reported to increase the number of serious illness conversations made by oncologists.^16^ In contrast, the adoption of an AI system for identifying atrial fibrillation patients at high risk of stroke failed to increase new anticoagulant prescriptions.^17^ These studies explore the potential for AI predictions to inform clinicians’ judgment collaboratively.

### What models are being deployed?

Trials predominantly evaluated deep learning systems for medical imaging (57/84, 67·9%). Notably, the medical imaging systems under evaluation were predominantly video-based (40/57, 70·2%) rather than image-based (17/57, 29·8%). This effect was primarily driven by the large number of endoscopy trials (32/40, 80·0%). Outside of imaging, AI systems operated on structured data, such as from the EHR (14/27, 51·9%), waveform data (10/27, 37·0%), and free text (3/27, 11·1%). These systems use a mix of decision trees (6/27, 22·2%), neural networks (2/27, 7·4%), reinforcement learning (2/27, 7·4%), Case-based reasoning (2/27, 7·4%), Bayesian classifiers (1/27, 3·7%), and unspecified machine learning (14/27, 51·9%).

Most systems operating on medical imaging (50/57, 87·7%) were evaluated in an assistive setup with a clinician, whereas models based on structured data tended to be compared against routine care (12/14, 85·7%). Models were developed primarily in industry (47/84, 56·0%) followed by academia (33/84, 39·3%), with the remaining four models having mixed or unstated origins.

### What are the trials’ findings?

**Table 2** summarizes the distribution of results and group comparisons. From the total of 84 trials, 79 attempted to demonstrate improvement while five used non-inferiority designs. Most of the trials that aimed to demonstrate improvement have reported a positive result for their primary endpoint (64/79, 81·0%). These trials principally noted improvements for AI-assisted clinicians compared to unassisted clinicians (47/64, 73·4%), and AI systems compared to routine care (14/64, 21·9%), with only three trials reporting superior performance from standalone AI to clinicians.

**Table 2.** Tabulated summary of primary endpoint results and group comparisons made for randomized controlled trials of AI in clinical practice.

|  | Primary Result |  |  |  |  |
| --- | --- | --- | --- | --- | --- |
| Comparison | Significant improvement | No significant improvement | Demonstrated noninferiority | Significant deterioration | Grand Total |
| AI vs Clinician | 3 | 1 | 3 | 1 | 8 |
| AI vs routine care | 14 | 4 | .. | .. | 18 |
| Assisted vs Unassisted | 47 | 9 | 2 | .. | 58 |
| Grand Total | 64 | 14 | 5 | 1 | 84 |

The five trials with non-inferiority designs established that there was no significant difference between standalone AI and clinicians (three trials) and between assisted unassisted clinicians (two trials).^8,12,18–20^ Hence, a total of 69 trials out of 84 (82·1%) have reported a positive result for their primary endpoint. A similar success rate was observed for the gastroenterology subset, with 27 of the 35 trials reporting significant improvement and one trial demonstrating noninferiority for an overall 80·0% success rate.

On the other hand, RCTs with a negative result for their primary endpoint include nine trials that failed to show an improvement of assisted clinicians compared to unassisted clinicians, four trials that failed to show an improvement of AI over routine care, and one trial that failed to show an improvement of standalone AI over clinicians. Additionally, one trial reported standalone AI to have significantly worse performance than clinicians. However, eight of these 15 trials reported a significant improvement for a secondary endpoint.^21–28^

## DISCUSSION

The scoping evaluation of AI RCTs reveals several noteworthy trends and implications for the development and implementation of AI in clinical practice. The distribution of trials across fields and locations highlights a concentration of AI applications in gastroenterology, radiology, surgery, and cardiology. The geographical distribution of trials reveals a dominance of single-country studies, with the US leading the way followed by China. This suggests a need for more international collaboration and multi-center trials to ensure the generalizability of AI systems across various populations and healthcare systems. Moreover, the preponderance of single-center trials implies a potential limitation in the generalizability of the results, which calls for an increased emphasis on multi-center trials in the future.

The deployment of deep learning systems for medical imaging, particularly video-based systems, is a prevalent trend in AI applications evaluated in RCTs. This is evident in the significant number of trials assessing video-based gastroenterology interventions, in contrast to the dominance of image-based radiology algorithms in academic literature and regulatory clearances.^29–32^ This trend appears to be driven by a few groups that account for most video-based gastroenterology trials, indicating that the field of clinical AI trials is still relatively homogeneous in terms of investigators, trial design, and outcome measures. Systems using structured data such as EHRs and waveform data, on the other hand, have employed a mix of decision trees, neural networks, reinforcement learning, and other machine learning techniques. This variety of models and data sources demonstrates the adaptability of AI to address different healthcare challenges. More research is needed to evaluate the impact of AI systems that incorporate clinical context (multiple modalities) or clinical priors (multiple timepoints) into their decision making, as these factors are critical to many clinical tasks.^33,34^

The majority of published RCTs for AI in clinical practice have had positive outcomes for their primary endpoints (69/84, 82·1%). This is a notably high success rate as compared to success rates found by both historical reviews of RCTs for medical interventions and recent reviews of RCTs for AI in healthcare.^35–38^ This disparity with recent reviews can be attributed to our definitions of AI and clinical practice, which led to the exclusion of studies lacking clinical integrations and non-linear AI, as well as our updated search of both trials and publication databases, which led to the inclusion of several new and previously overlooked trials.^36–39^ While such a high success rate lends credibility to the promise of clinical AI, this should be tempered by recognition of the nascency of the field and the likeliness of publication bias.

Most trials evaluated interventions on diagnostic accuracy-related outcomes. While such trials offer convincing evidence of the prospective technical performance of clinical AI, this may not accurately reflect its overall impact on patient care, as high sensitivity and specificity do not necessarily translate to improved patient outcomes. For example, a recent systematic review of 21 colonoscopy trials found that, while AI assistance helped increase polyp detection, it did not yield significant increases in the detection of clinically critical advanced adenomas. ^40^ Some trials have assessed the impact of AI on care management quality metrics, patient behavior and symptoms, and clinical decision-making. These diverse outcome measures reflect the various ways in which AI can influence clinical practice, from improving care quality to enhancing patient experience and informing clinical judgment. To better assess the true value of AI algorithms in healthcare, it is crucial to incorporate clinically meaningful endpoints such as survival, symptoms, and need for treatment.

In conclusion, the existing landscape of RCTs on AI in clinical practice demonstrates an expanding interest in applying AI across a range of fields and locations. Most trials report positive outcomes, highlighting AI’s potential to potentially enhance care management, patient behavior and symptoms, and clinical decision-making. To understand AI’s true impact and limitations more comprehensively in healthcare, further research is essential, including a focus on multi-center trials and the incorporation of diverse outcome measures.

## Authors’ contributions

RH and PR conceptualized the study. JPAI, PR, and EJT supervised the study. RH, JPAI, and PR contributed to the design of the study. RH, JNA, and PR carried out the screening of the search results and did the data extraction. RH drafted the manuscript. ZS contributed to the presentation of data. All authors had access to the data and critically revised and edited the manuscript.

## Declaration of interests

EJT receives funding from NCATS/NIH grant UL1TR002550. RH is an employee of Quadrant Health, outside of the submitted work. JNA is an employee of Rad AI Inc, outside of the submitted work. All other authors declare no competing interests.

## Data Availability

Search strategy and raw data will be made available in the appendix upon publication.

## Acknowledgements

RH pursued this work while supported by the Summer Institute for Biomedical Informatics at Harvard Medical School.

## Notes

### Funding Statement

This study did not receive any specific funding.

